# COVID 19 in Bangladesh: Assumption of possible infection and death

**DOI:** 10.1101/2020.05.17.20104562

**Authors:** Sadik Hasan Shuvo

## Abstract

It is a painful job to predict the death of people. But sometimes it is important to predict the future and concern the government. A furious future is waiting for Bangladesh.

**Objective:** Objective of the study is to assume the number of positive case and death till 30^th^ December, 2020 in Bangladesh.

**Study design:** This study was designed with systematic review and data analysis.

**Method:** The study was completed by analyzing data available on website. First COVID 19 case in Bangladesh was identified on 8^th^ March. Analyzing the data increasing rate/common ratio of infection and death has been identified. Then this common ratio has been used in the formula of multiplication series (at decreasing rate). Data of China, Iran, Italy and the USA was also analyzed to assume how the death and case number increased. Social issues of Bangladesh were also analyzed. Considering all these the assumption was made.

**Result:** It has been assumed that by the 43^rd^ week (on 30^th^ December, 2020) of first identification the total case can be 15640747 and total death can be 638769 by 30 December, 2020. As this is an assumption this can be true, partially true or false. But the base of assumption is strong enough so the possibility of being true or nearly true is higher.

**Policy Suggestion:** Government should choose properly one between two options. Either government should declare curfew or let people lead normal life for the purpose of herd immunity. A very weak lockdown for a long time doesn’t make any sense.

## 1.1. Introduction

Bangladesh is one of the most vulnerable countries to COVID 19 in the world. Till 20^th^ May 386 people died and total case is 26738^i^. Just 10 week ago on 8^th^ March first COVID 19 patient was identified and first death case was on 18^th^ March^ii^ Since then the number of both positive case and death is increasing. Within a very few time the virus is showing its devastating behavior in this country. The government has a lot to do still now but if they don’t take proper steps what will happen? This article shows an assumption about the upcoming future. This assumption is not just a mere prediction. It is a mathematical certainty.

## 1.2. Significance of the Study

This study will be helpful for the government and the policy maker. Still neither the people nor the government is concern enough about the virus. This study will show them the future with date. This will appear as live data to them and it will help them to think carefully and be more concern.

## 1.3. Research Questions

This study tried to find out the answer of the following questions.

- What is the increasing rate of positive case and death?
- What is waiting for the future?
- How many people will be affected and how many will die?
- How the positive case and death number will be increased?

## 1.4. Methodology of the Study

The following sequence shows the incensement of positive case in every week in Bangladesh.

3+14+39+54+218+1231+3772+7103+11719+17822+26738……………………….

The following sequence shows the incensement of death number in every week in Bangladesh.

1+5+6+20+50+120+163+186+269+386……………………….

No available arithmetic formula works here as there is no common ratio or common difference of the increasing number. So author designed a formula to find out the common ratio. Then the common ratio has been used in the formula of multiplication series to find out the possible number of positive case and death. The whole process is described here.

**First Part:** R = (nearest ratios)/n
**Second Part:** *n-th* term = N*R^n-1 (Formula of multiplication series)

Simply the sequence is built by multiplying the previous number with common ratio

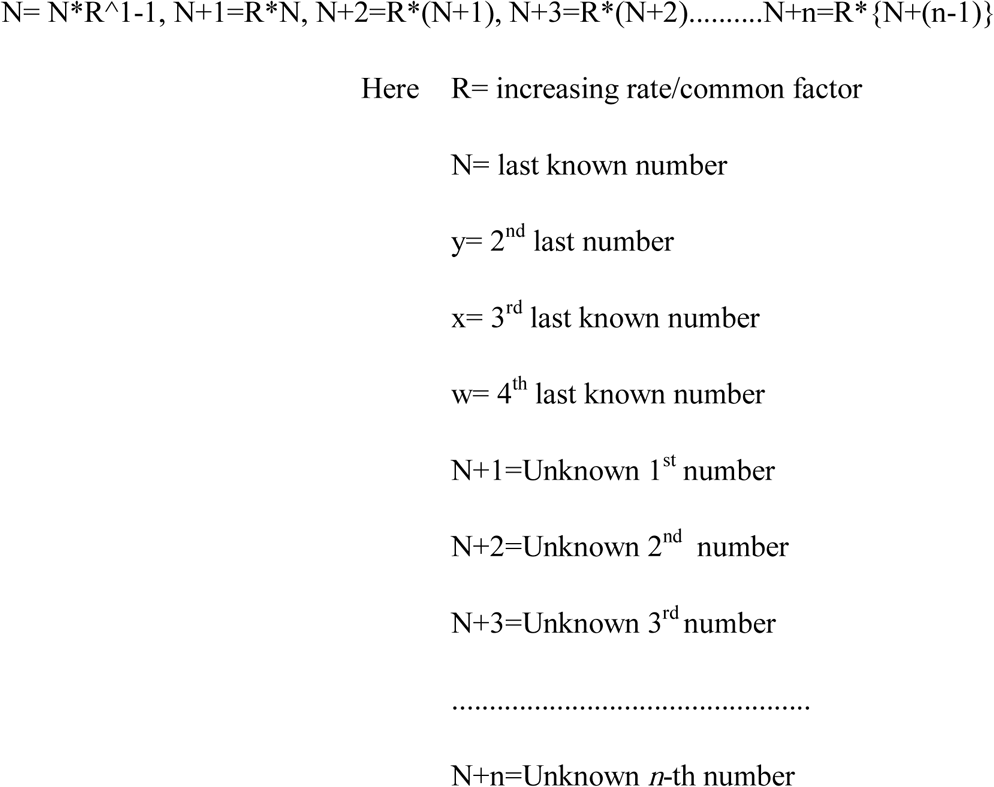

### 1.4.1. Explanation of R= increasing rate/common ratio

In multiplication series R or common ratio (increasing rate) is determined by a formula, **r = (n+1)/n**. simply it divides each term by the previous term. But in this case there is no common ratio as it is increasing in a natural way. In this case mean of the nearest ratios has been considered as “R”

To find out the common ratio the mean formula has been used here. Increasing rate of last four weeks has been taken. Then the mean of the nearest rates of three weeks have been calculated and the result has been considered as the common ratio or increasing rate, r.

Data of the last three weeks have been taken as these weeks show a stable difference. For the COVID 19 positive cases the ratio of last weeks’ are 1.64, 1.52 and 1.50 which are very close to each other. But the ratios of previous weeks are: 4.7, 2.8, 1.4, 4, 1.9 and 5.6. The ratios in these weeks are so much fluctuating. So these ratios have been ignored in mean calculation.

Similarity has been found in the ratio of death number. For the death number the ratios of three weeks’ among last four are 1.4, 1.4, and 1.4 which are same. But the ratios of previous weeks are: 5, 1.2, 3.3, 1.1 and 6. The ratios in these weeks are so much fluctuating. So these ratios have been ignored in mean calculation.

It has been found that the common ratio decreases in other countries in every month so the ratio has also been used in the formula with a decreasing rate. The rate was decreased considering the current rate and the socio economic condition of Bangladesh.

### 1.4.2. Calculation of Positive case

**Mean ratio**,

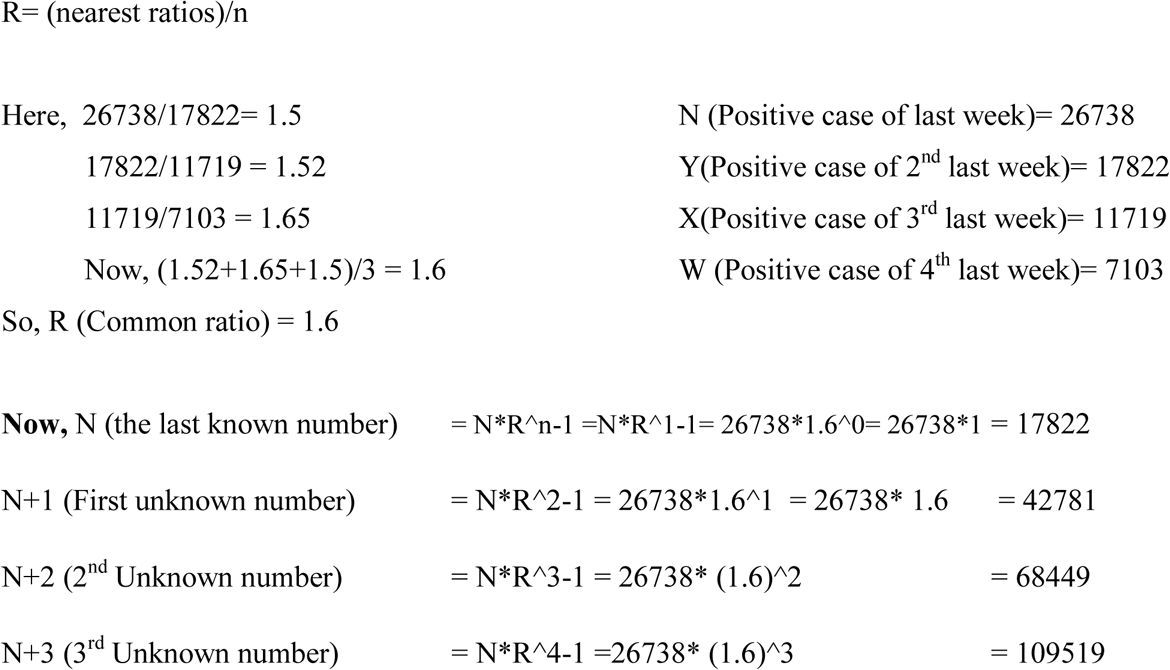

The possible positive cases have been calculated up to 30^th^ December. (Shown in table 1 and diagram 1)

**Table 1:** Series of COVID 19 positive case.

| Week | Date (2020) | Total Positive case | Increasing rate | Week | Date (2020) | Total Positive case | Increasing rate |
| --- | --- | --- | --- | --- | --- | --- | --- |
| week 1 | 11-Mar | 3 |  | Week 23 | 12-Aug | 2171358 | 1.20 |
| week2 | 18-Mar | 14 | 4.67 | Week 24 | 19-Aug | 2605630 | 1.20 |
| week3 | 25-Mar | 39 | 2.79 | Week 25 | 26-Aug | 3126756 | 1.20 |
| week4 | 1-Apr | 54 | 1.38 | Week 26 | 2-Sep | 3752107 | 1.20 |
| week5 | 8-Apr | 218 | 4.04 | Week 27 | 9-Sep | 4127318 | 1.10 |
| week6 | 15-Apr | 1231 | 5.65 | Week 28 | 16-Sep | 4540050 | 1.10 |
| week7 | 22-Apr | 3772 | 3.06 | Week 29 | 23-Sep | 4994055 | 1.10 |
| week8 | 29-Apr | 7103 | 1.88 | Week 30 | 30-Sep | 5493460 | 1.10 |
| week9 | 6-May | 11719 | 1.65 | Week 31 | 7-Oct | 6042806 | 1.10 |
| Week 10 | 13-May | 17822 | 1.52 | Week 32 | 14-Oct | 6586659 | 1.09 |
| week 11 | 20-May | 26738 | 1.50 | Week 33 | 21-Oct | 7179458 | 1.09 |
| week12 | 27-May | 42781 | 1.60 | Week 34 | 28-Oct | 7825609 | 1.09 |
| week13 | 3-Jun | 68449 | 1.60 | Week 35 | 4-Nov | 8529914 | 1.09 |
| week14 | 10-Jun | 109519 | 1.60 | Week 36 | 11-Nov | 9297606 | 1.09 |
| week15 | 17-Jun | 175230 | 1.60 | Week 37 | 18-Nov | 10041415 | 1.08 |
| week16 | 24-Jun | 280368 | 1.60 | Week 38 | 25-Nov | 10844728 | 1.08 |
| week17 | 1-Jul | 392516 | 1.40 | Week 39 | 2-Dec | 11712306 | 1.08 |
| week18 | 8-Jul | 549522 | 1.40 | Week 40 | 9-Dec | 12649291 | 1.08 |
| week19 | 15-Jul | 769330 | 1.40 | Week 41 | 16-Dec | 13661234 | 1.08 |
| week20 | 22-Jul | 1077063 | 1.40 | Week 42 | 23-Dec | 14617520 | 1.07 |
| Week 21 | 29-Jul | 1507888 | 1.40 | Week 43 | 30-Dec | 15640747 | 1.07 |
| Week 22 | 5-Aug | 1809465 | 1.20 |  |  |  |  |

**Diagram 1:**
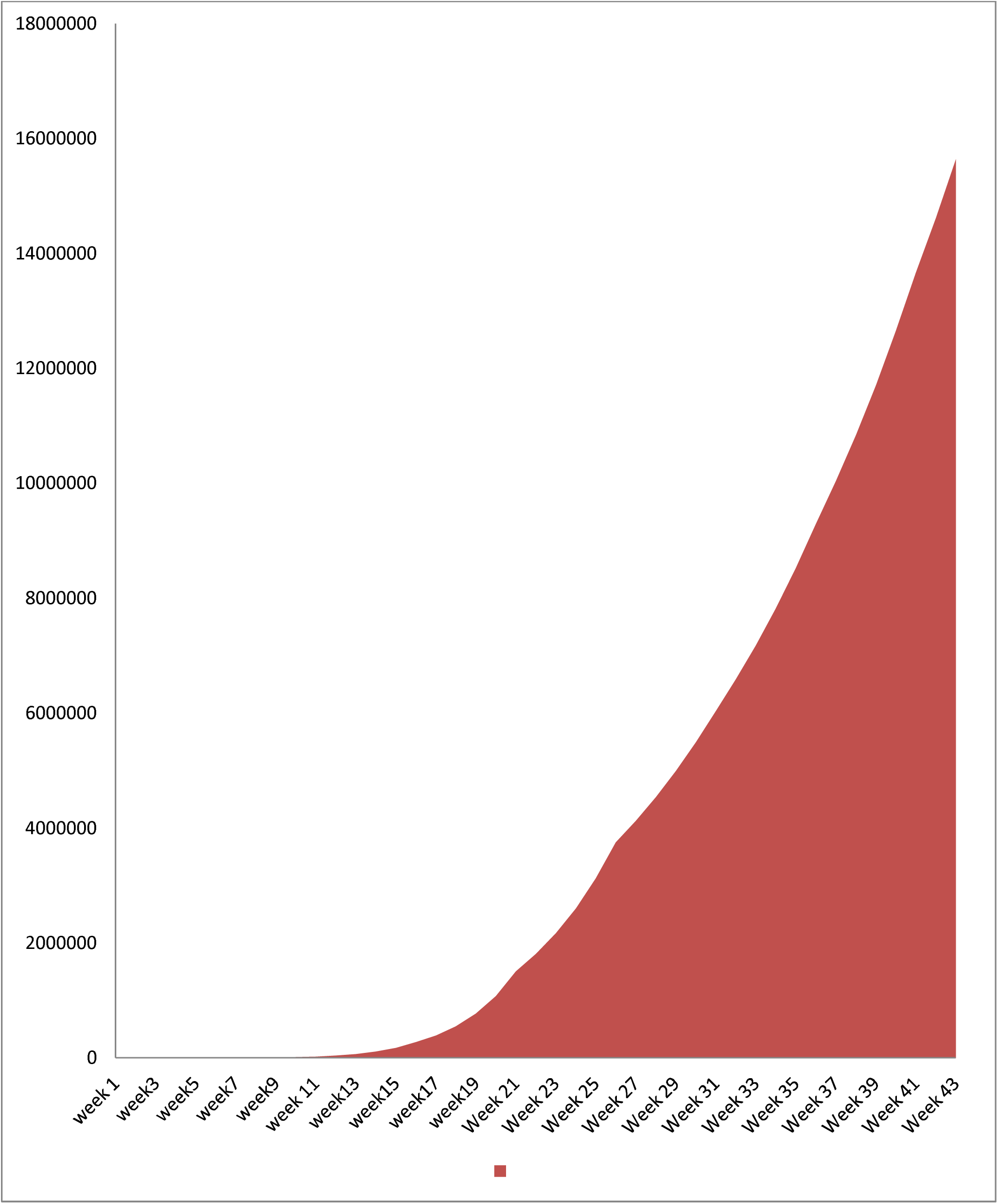
Series of COVID 19 positive case.

### 1.4.3. Calculation of death number

The death number was identified in the same way that of possible positive case identification.

**Mean ratio,**

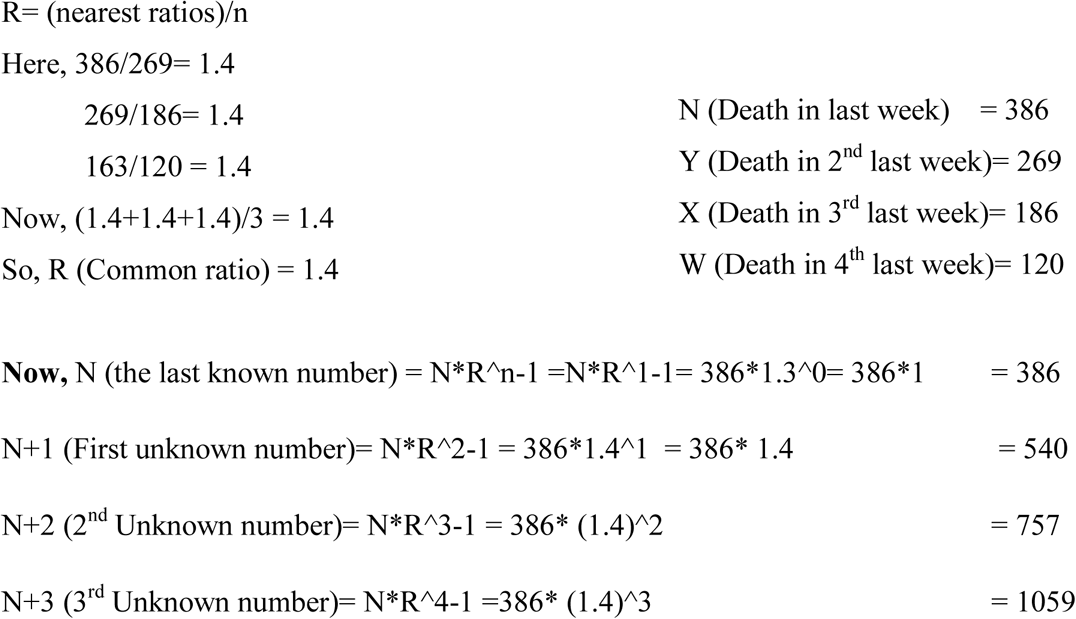

The possible deaths have been calculated up to 30^th^ December. (Shown in table 2 and diagram 2)

**Table 2:** Series of Death of COVID 19.

| **Week** | **Date (2020)** | **Total Death** | **Increasing rate** | **Week** | **Date (2020)** | **Total Death** | **Increasing rate** |
| --- | --- | --- | --- | --- | --- | --- | --- |
| week 1 | 18-Mar | 1 |  | Week 22 | 12-Aug | 15732 | 1.30 |
| weed 2 | 25-Mar | 5 | 5.00 | Week 23 | 19-Aug | 20452 | 1.30 |
| week3 | 1-Apr | 6 | 1.20 | Week 24 | 26-Aug | 26587 | 1.30 |
| week4 | 8-Apr | 20 | 3.33 | Week 25 | 2-Sep | 34563 | 1.30 |
| week5 | 15-Apr | 50 | 2.50 | Week 26 | 9-Sep | 43204 | 1.25 |
| week6 | 22-Apr | 120 | 2.40 | Week 27 | 16-Sep | 54005 | 1.25 |
| week7 | 29-Apr | 163 | 1.36 | Week 28 | 23-Sep | 67506 | 1.25 |
| week8 | 6-May | 186 | 1.14 | Week 29 | 30-Sep | 84383 | 1.25 |
| week9 | 13-May | 269 | 1.45 | Week 30 | 7-Oct | 105478 | 1.25 |
| Week 10 | 20-May | 386 | 1.43 | Week 31 | 14-Oct | 126574 | 1.20 |
| week 11 | 27-May | 540 | 1.40 | Week 32 | 21-Oct | 151889 | 1.20 |
| week12 | 3-Jun | 757 | 1.40 | Week 33 | 28-Oct | 182267 | 1.20 |
| week13 | 10-Jun | 1059 | 1.40 | Week 34 | 4-Nov | 218720 | 1.20 |
| week14 | 17-Jun | 1483 | 1.40 | Week 35 | 11-Nov | 262464 | 1.20 |
| week15 | 24-Jun | 2076 | 1.40 | Week 36 | 18-Nov | 301833 | 1.15 |
| week16 | 1-Jul | 2803 | 1.35 | Week 37 | 25-Nov | 347108 | 1.15 |
| week17 | 8-Jul | 3784 | 1.35 | Week 38 | 2-Dec | 399175 | 1.15 |
| week18 | 15-Jul | 5108 | 1.35 | Week 39 | 9-Dec | 459051 | 1.15 |
| week19 | 22-Jul | 6895 | 1.35 | Week 40 | 16-Dec | 527908 | 1.15 |
| week20 | 29-Jul | 9309 | 1.35 | Week 41 | 23-Dec | 580699 | 1.10 |
| Week 21 | 5-Aug | 12102 | 1.30 | Week 42 | 30-Dec | 638769 | 1.10 |

**Diagram 2:**
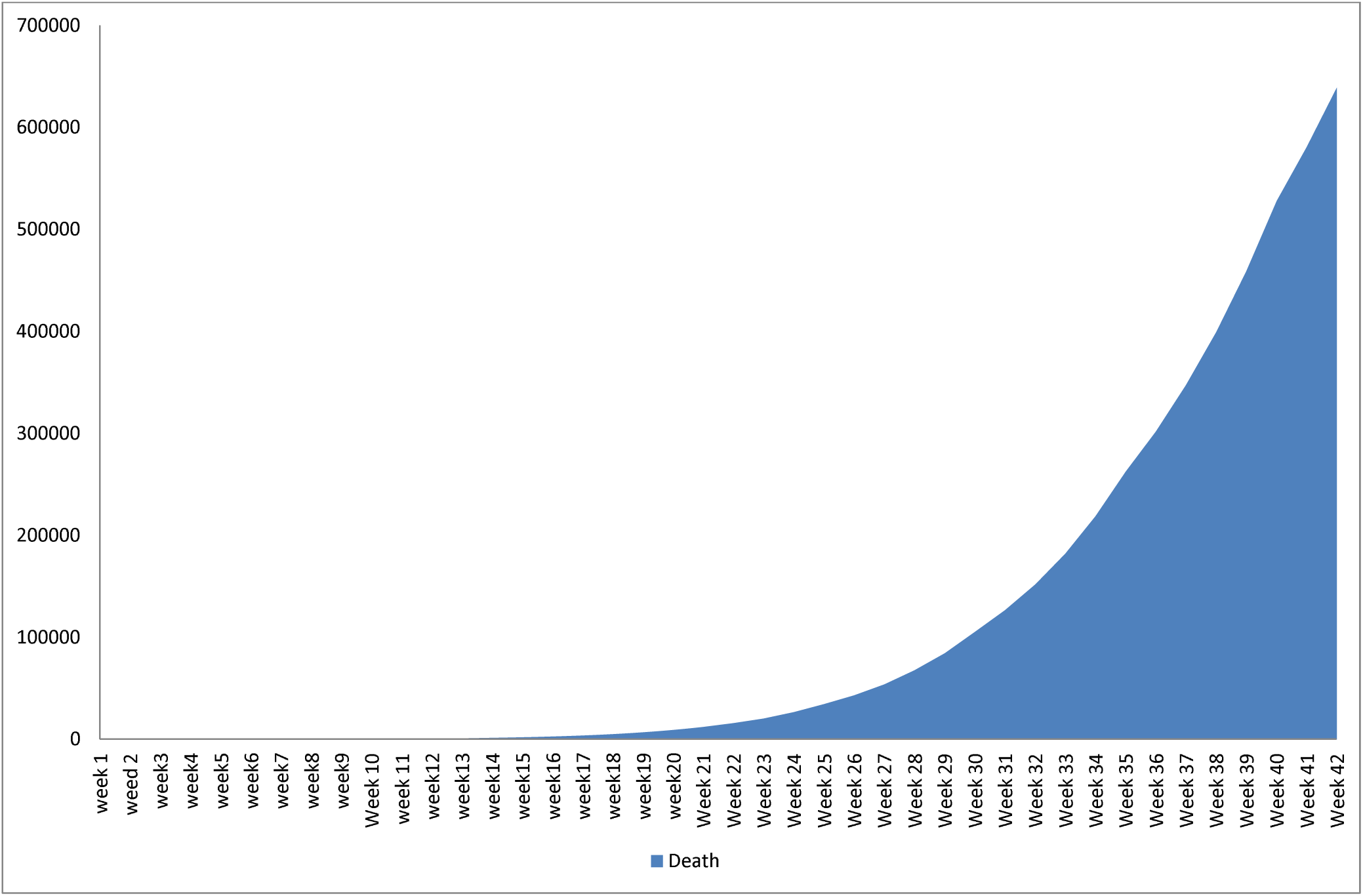
Series of Death of COVID 19.

### 1.4.4. Changes in common ratio

Data of five countries (China^iii^, Iran^iv^, Italy^v^, USA^vi^ and Bangladesh^vii^) has been analyzed. It has been found that both death rate and rate of positive cases are increasing at a decreasing rate (showed in figure 3 and 4). From this it can be assumed that the rate of positive case and death in Bangladesh will be increased at a decreasing rate. The current data shows that in the ninth week the rates in Bangladesh are higher than other four countries in both death and positive cases. So it can be assumed that though the rates will be decreased it will not be like other countries. Sometimes it increases also. Increasing and decreasing depend on many things such as health care, lockdown, awareness etc. Analyzing all these it can be predicted that both death rate and the rate of infection will be decreased in every five weeks. The decreasing rates were determined observing the rates of other countries and the overall condition of Bangladesh.

**Figure 3:**
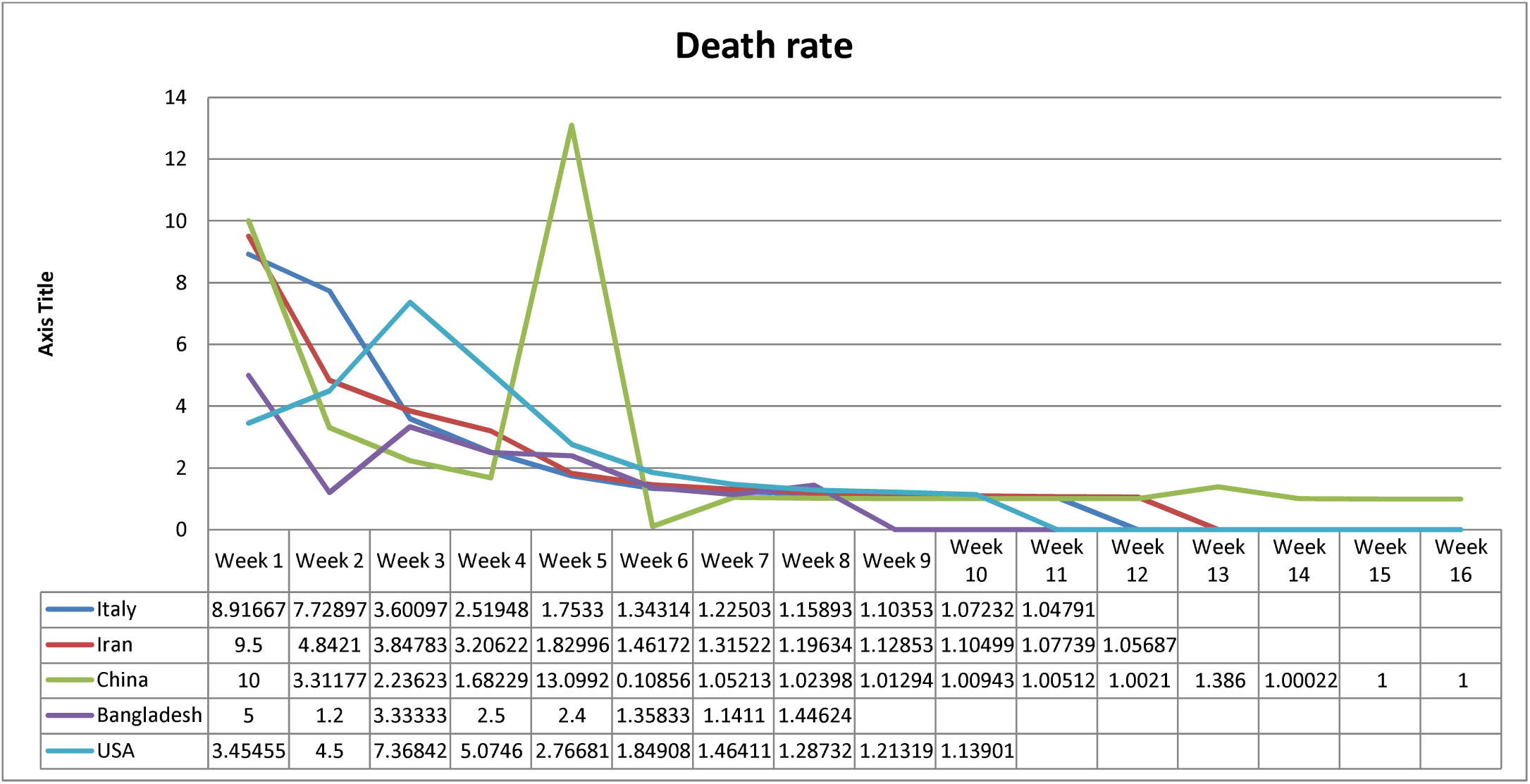
Death rate of Italy, Iran, China, Bangladesh and the USA

**Figure 4:**
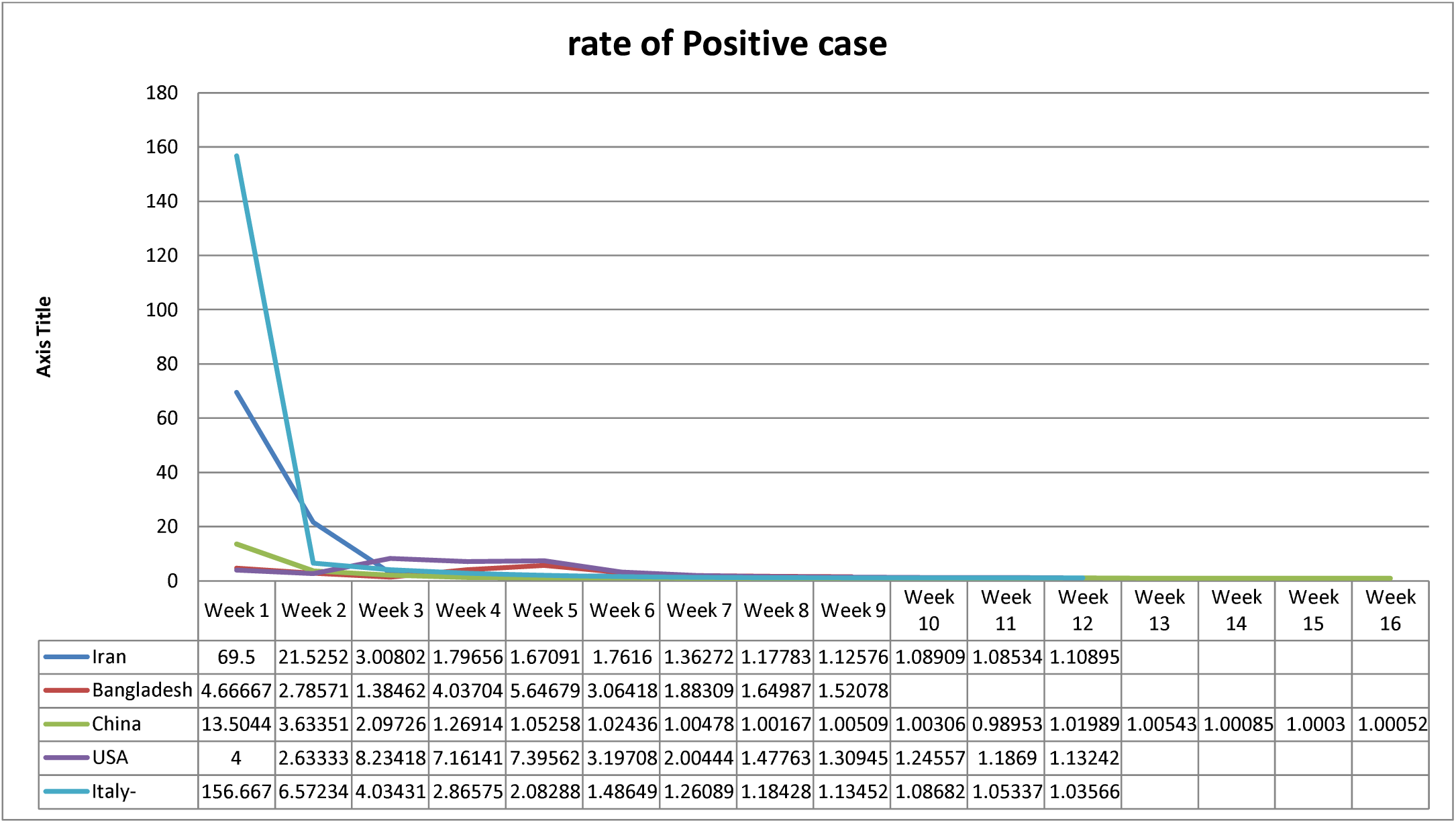
Rate of positive case of Italy, Iran, China, Bangladesh and the USA

## 2. Weakness of the Study

It is not possible to predict the future perfectly. Based on data it can only be assumed. Assumption can be true, partially true or false. But if assumption is based on data, mathematical formula, social and other context then the possibility of perfection becomes higher. In this study data of five countries has been analyzed, formula of multiplication series has been applied and the health, social, political context of Bangladesh has been taken into consideration. So the possibility of being true of this assumption is higher but not 100%.

## 3. Policy suggestion

Government should impose stick lockdown (curfew) or it should go for herd immunity. But in Bangladesh lockdown is going on and amid of this garments factories and markets are opened. The number of both positive case and death is increasing rapidly. Government should take immediate action to tackle the situation.

## Data Availability

Yes, data is available

## Authors’ Statement

There was no funding for the research. There is no competing interest also. Ethical approval was not taken as this study is based on secondary data.

## Footnotes

i IEDCR (15th May, 2020) Covid-19 Status Bangladesh, available at: https://cutt.ly/byWORph accessed on: 15th May, 2020

ii Worldometer (15th May, 2020) Bangladesh, available at: https://cutt.ly/tyWOOdQ accessed on: 15th May, 2020

iii Worldometer (15th May, 2020) China, available at: https://cutt.ly/fyEPndS accessed on: 15th May, 2020

iv Worldometer (15th May, 2020) Iran, available at: https://cutt.ly/iyEPbcl accessed on: 15th May, 2020

v Worldometer (15th May, 2020) Italy, available at: https://cutt.ly/4yEPbuA accessed on: 15th May, 2020

vi Worldometer (15th May, 2020) USA, available at: https://cutt.ly/syEPvYc accessed on: 15th May, 2020

vii Ibid

